# A Novel Coach-Approach to Clinical Faculty Mentoring and the UW Department of Medicine Clinical Faculty Development Program

**DOI:** 10.1101/2024.03.21.24304694

**Authors:** James D Alstott, Chariti Gent, Christine Fabian Bell, Daniel R Marlin, Anthony Hernandez, Esther Schulman, Sharon Gehl, Lynn M Schnapp, James H Stein

**Author notes:** **Corresponding Author:** James H. Stein, MD, University of Wisconsin School of Medicine and Public Health 600 Highland Avenue, Room H4/520 CSC (MC 3248), Madison, Wisconsin 53792.

## Abstract

**Background:** Clinical faculty at academic health centers may benefit from specific mentorship and proficiencies that are distinct from those on research tracks. We describe the creation, activities, and one-year impact of a faculty development program that included novel professional coaching training (the Clinical Faculty Mentoring Program, CFMP) which was supplemented by skills- and knowledge-building activities (the Clinical Faculty Development Series, CFDS).

**Methods:** The goals and components of the CFMP and CFDS are described in detail. A mixed methods evaluation plan guided collection of confidential survey and interview data before and after the first year of these activities. We used paired t-tests to identify statistically significant changes.

**Results:** The 43 clinical mentors reported significant gains in job satisfaction, teaching attitudes, knowledge of mentorship competencies, and confidence with coaching skills for mentorship (all p<0.05). Of mentor respondents, 88% found the coach approach to mentoring program to be “very” or “somewhat” helpful. Coaching behavioral domains with the greatest evidence of improvement were supporting the mentee to integrate new awareness, insight, learning into their worldview and behaviors (p=0.0503) and managing time and focus of mentoring sessions (p=0.022). All 37 mentees had at least one meeting with a mentor (100%). Over 9 months, 39 virtual CFDS sessions had an average participation of 38 participants (range 22-59). A majority of surveyed faculty (>55%) agreed or strongly agreed the CFDS sessions provided valuable opportunities for skills development with teaching, leadership, wellness, diversity, equity, inclusion, and promotion.

**Conclusions:** Among clinical mentors, our novel coach approach to clinical faculty mentoring and skill-building had favorable effects on job satisfaction, knowledge of mentorship competencies, and confidence in coaching skills. Outcomes from the clinical faculty development series supported the mentoring program outcomes. Longitudinal follow-up is needed to determine how this program will impact mentees.

## Background

Engaged faculty with a strong sense of professional fulfillment and organizational value are vital to sustaining and growing academic health centers’ tripartite mission of clinical service, education, and research. However, early career faculty may lack the understanding and skills to navigate successful careers at an academic health center and their professional identity formation and fulfillment increasingly are threatened by greater clinical and administrative demands, burnout, and balancing work-life integration. These issues are amplified for women and persons underrepresented in medicine minority groups (URiM) in part due to a perception of low institutional inclusion and promotion to leadership roles^1,2^.

Faculty development programs at academic health centers have assumed responsibility for advancing faculty towards promotion, supporting mentorship, creating collaborative networks, and fostering education, research, and additional professional skills^3,4^, however most of the literature regarding the effectiveness of mentoring academic health centers has focused on scientific researchers, not clinicians. Some data suggest that faculty development programs may increase career satisfaction and engagement, utilization and satisfaction with mentorship opportunities, research productivity, and promotion rates^5–10^. Indeed, faculty who participate in professional development are more fulfilled, productive, and are less likely to leave their institution^5,11–14,15^. Mentoring programs have been identified as particularly impactful at promoting faculty vitality; however, mid-career and senior clinical faculty may not have the knowledge base and mentoring skills to provide impactful guidance^9,16^ and the optimal approach to mentor training and its effects on the mentor’s career development and professional vitality are not known.

To address the needs of clinicians in the University of Wisconsin (UW) Department of Medicine (DOM), we developed and implemented a faculty development program that had two components: a Clinical Faculty Mentoring Program (CFMP) which used a novel coaching approach to train faculty mentors, and a Clinical Faculty Development Series (CFDS), with a unique focus on topics of specific interest to clinicians (Table 1). Traditional mentoring programs are mentor-driven with the mentor functioning as a problem-solver who provides advice and guidance to mentees regarding career goals and challenges. The CFMP used coaching principles to teach clinician mentors how to focus on mentee behaviors and how to foster their mentee’s self-awareness and growth using principles from positive psychology and motivational interviewing. The primary aim of this program was to provide clinicians with robust experiences that cultivate skill-building, mentorship, and opportunities to enhance professional satisfaction and engagement with the promotion process. The CFDS supported this training and was directed at all clinical faculty in the UW DOM. The long-term goals of these programs are to increase clinician vitality, attenuate burnout and physician distress, improve the DOM’s climate, and increase faculty retention. In this paper, we describe the activities and impact of the first year of the CFMP and CFDS with particular focus on the mentors who completed the novel coach approach to clinical mentoring.

**Table 1:**
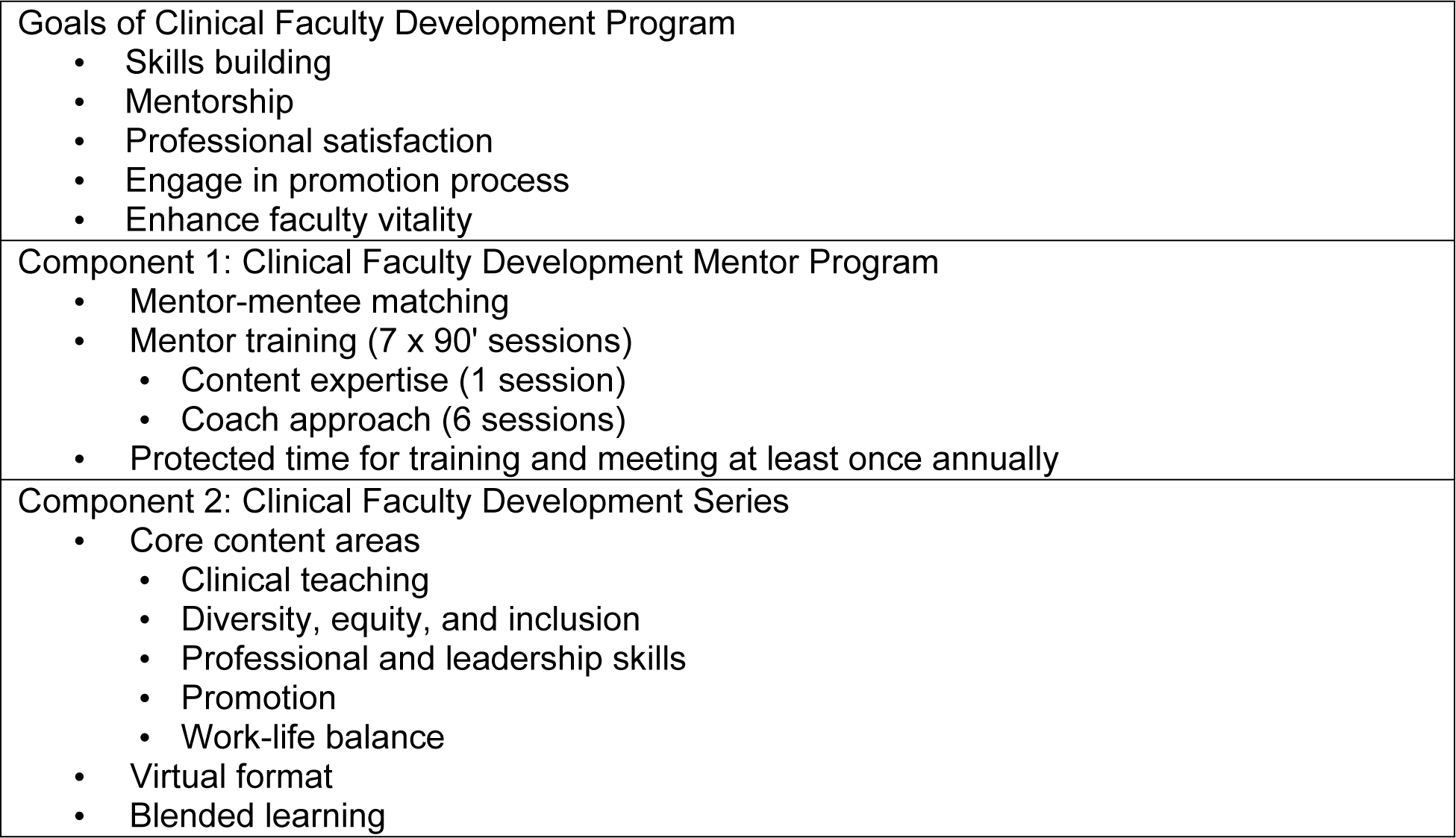
University of Wisconsin Department of Medicine Clinical Faculty Development Program - Structure and Activities.

|  |
| --- |
| Goals of Clinical Faculty Development Program<br>• Skills building<br>• Mentorship<br>• Professional satisfaction<br>• Engage in promotion process<br>• Enhance faculty vitality |
| Component 1: Clinical Faculty Development Mentor Program<br>• Mentor-mentee matching<br>• Mentor training (7 x 90' sessions)<br>• Content expertise (1 session)<br>• Coach approach (6 sessions)<br>• Protected time for training and meeting at least once annually |
| Component 2: Clinical Faculty Development Series<br>• Core content areas<br>• Clinical teaching<br>• Diversity, equity, and inclusion<br>• Professional and leadership skills<br>• Promotion<br>• Work-life balance<br>• Virtual format<br>• Blended learning |

## Methods

The UW-Madison Health Sciences Human Subjects Committee (the institutional review board [IRB] for the UW School of Medicine and Public Health) determined that our evaluations did not meet the definition of human subjects research. They determined that the activities and analyses described in this report were considered quality assurance and declined to review them or to request completion of IRB-approved consent forms. Participation in all surveys was fully anonymous, without any records or identifiers of who did and did not participate. Informed consent for participation in the interviews was provided orally to Wisconsin Center for Education Research staff. All data, including who chose to participate in these interviews, were analyzed anonymously. No minors or prisoners participated in this study.

### Setting

The UW DOM is comprised of 446 faculty in 11 divisions. This mentoring program was designed for clinical faculty who spend most of their time in direct patient care; excellence in clinical practice is the primary goal for their academic promotion with consideration of significant accomplishments in teaching and service. Most clinical track faculty in the UW DOM are at the rank of Assistant Clinical Professor (56%), followed by Associate Clinical Professors (30%) and Clinical Professors (14%). The mean (standard deviation) age for clinical faculty by rank are 42 (10.1, range 28-75) years for Assistant Clinical Professors, 49 (8.3, range 36-74) years for Associate Clinical Professors, and 55 (6.1, range 45-68) years for Clinical Professors. Females comprise 45% of clinical track faculty. The UW does not disclose race/ethnicity distribution of faculty, though we allowed voluntary disclosure of race/ethnicity from participants in our program.

### Clinical Faculty Mentoring Program (Table 1)

The mentorship component of the CFMP was created following the Science of Effective Mentorship by the National Academies of Sciences, Engineering, and Medicine: a professional, working alliance in which individuals work together over time to support the personal and professional growth, development, and success of the relational partners through the provision of career and psychosocial support^17^.

Assistant Clinical Professors who had joined the UW DOM after July 2020 were required by departmental promotion guidelines to participate in the CFMP. The first step was identifying mentors. Mentors were recruited by email to all Associate or full Clinical Professors in the DOM. Division Heads also encouraged their faculty participation at Division meetings. Next, mentors and mentees completed surveys that indicated their professional interests and preferences for mentor-mentee matching (*i.e*., same/different academic Division, gender identity, race/ethnicity, professional interests, non-professional interests; see Supplemental Text 1 and 2). After surveys were completed, a DOM staff member and the Vice Chair for Faculty Development matched mentees with mentors based on survey responses; requests regarding specialty, professional interests, and gender identity were prioritized and tentative matches. Then, mentors completed a structured training curriculum that consisted of seven, 90-minute virtual sessions from September 2022 through June, 2023. The first session covered three components of the mentoring knowledge base: promotion standards and processes; faculty well-being and institutional resources; and diversity, equity, and inclusion (DEI) resources.

The next six sessions used a novel “Coach Approach to Clinical Faculty Development” to deliver information about professional coaching competencies. Utilizing the International Coaching Federation’s (ICF) core competency framework and drawing from the UW’s ICF-accredited Certified Professional Coach program curriculum, each of the six sessions provided lessons in the foundational components of a coach approach to faculty mentoring^18^. Components included but were not limited to understanding the coaching mindset, designing the mentor-mentee relationship/alliance, and communicating effectively via powerful questioning and listening actively^19^. These sessions integrated hands-on and experiential exercises inside and outside of class for mentors to practice their newly acquired skills. Mentors were excused from clinical activities during their training sessions and to meet with their mentees. They were encouraged to meet with their mentees at least once in the first year of the program. Recommendations to start meeting were made after four of the seven training sessions were completed.

### Clinical Faculty Development Series (Table 1)

We simultaneously initiated a CFDS that provided weekly, one-hour learning sessions that focused on a wide variety of topics related to clinical faculty, including promotion, teaching, professional and leadership skills, work-life balance, and DEI (Supplemental Table 1: CFDS Session Titles and Categories). The CFDS was open to all clinical faculty in the UW DOM, not just participants in the CFMP. Each session provided a blended learning opportunity that usually included didactic and interactive components such as small group breakout sessions, “open mic” large group discussions, and role-playing/simulation activities. Content for each of the 39 sessions was provided by UW experts in each field. CFDS sessions occurred every Tuesday over the noon hour from September 2022 through June 2023 and were held virtually to maximize attendance of faculty working across various geographic sites in the UW DOM. CFDS lectures were recorded and uploaded with the permission of the presenter to an internal video lecture archive for faculty who could not attend to view asynchronously. The CFDS was promoted via institutional email to all UW DOM faculty, internal video DOM, websites, and DOM newsletter. Weekly reminder emails were sent to faculty with the upcoming week’s CFDS topic.

### Program Evaluations

The timeline of the CFMP activities and evaluations are shown in Figure 1. Professional staff from the Wisconsin Evaluation Collaborative at the Wisconsin Center for Education Research led the evaluation of the CFMP. A mixed methods evaluation plan guided collection of confidential survey and interview data. Qualtrics surveys were used to collect mentor and mentee baseline data before the program began and a post-survey that coincided with the end of the coach approach to mentoring sessions. Both pre- and post-surveys focused on understanding of promotion processes, satisfaction with workplace processes, and workplace climate. Mentor surveys included confidence in mentoring skills. After completion of the coaching training, mentors also were surveyed for impressions, utilization, and feedback. The data from the post-surveys are described below. Responses from mentors and mentees that completed both pre- and post-surveys were used to evaluate the effects of the CFMP on the outcomes. Additionally, six mentors and five mentees participated in confidential, semi-structured interviews over Zoom with experts from Wisconsin Center for Education Research about their experience with the CFMP. Interviews were audio recorded, transcribed, and thematically coded with NVivo software (Lumivero, Denver, CO).

**Figure 1.**
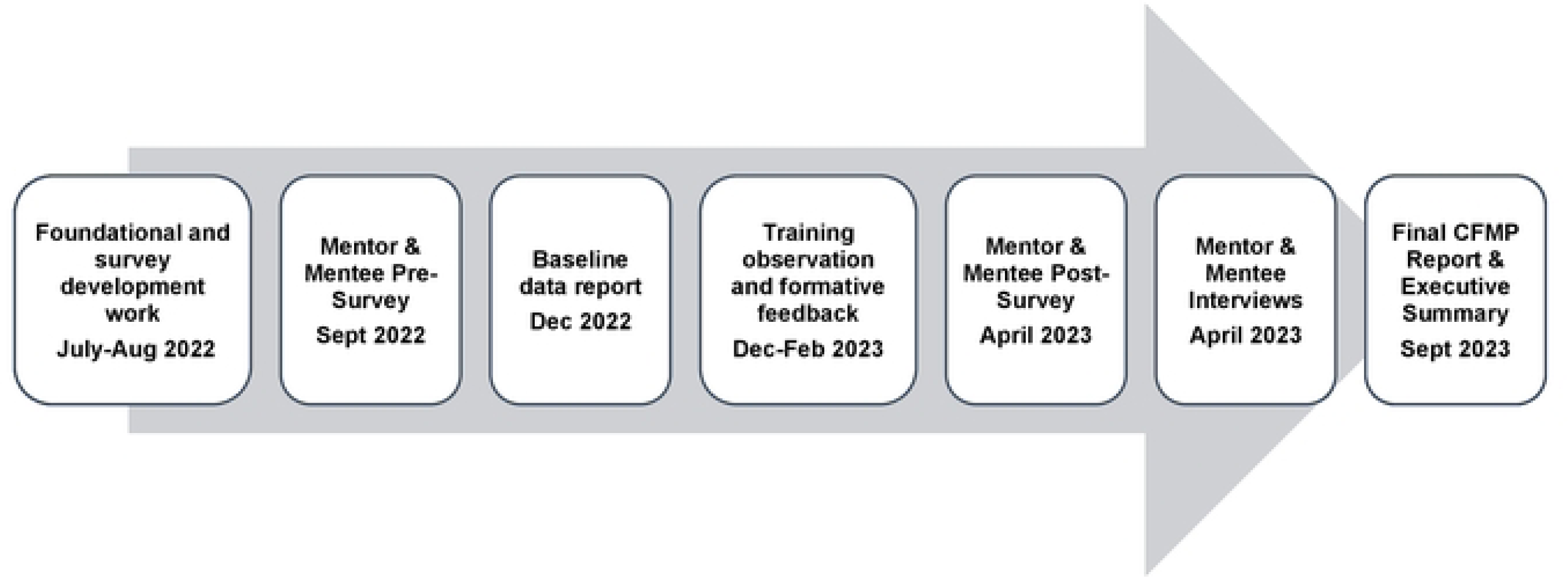
Timeline of Clinical Faculty Development Mentorship Program Activities and Evaluation Process.

Participant attendance data for the CFDS were obtained for each session. After the final session, Qualtrics surveys were used to elicit semi-quantitative feedback and qualitative responses from participating faculty. The question “Based on what you experienced as part of the CFDS, to what extent do you agree…” was used to assess whether the aims of the CFDS were successful. A 5-point Likert scale (1=strongly disagree to 5=strongly agree) was used for responses. Values are described as means and standard deviations. Paired t-tests were used to compare pre- to post-survey values among participants that completed both surveys. A p-value of <0.05 was considered statistically significant. P-values were not adjusted for multiple comparisons given the sample size; they were interpreted conservatively and in context.

## Results

### CFMP Participants

Response rates for participating CFMP faculty are shown in Table 2. Demographic data from the post-survey are in Table 3. Most mentors were Associate Clinical Professors and had been employed by the UW DOM for 6-19 years. A similar number of male and female mentors participated; among mentees there were more males and participants who preferred not to report their gender identity. Most mentors were white; mentees had more diverse URiM representation. Faculty from nine clinical divisions within the DOM participated in the mentoring program. Of post-survey respondents, 67% of mentors and 100% of mentees reported having had at least one mentor/mentee meeting and 28% of mentors reported meeting more frequently with their mentee, on either a monthly or quarterly basis. Of note, some mentors had more than one assigned mentee and some mentors had not yet been assigned a mentee by the end of the first year.

**Table 2:** Clinical Faculty Mentoring Program Survey Response Rates.

|  | **Pre-Survey**<br>**N (%)** | **Post-Survey**<br>**N (%)** | **Both Pre- and Post-Post Survey**<br>**N (%)** |
| --- | --- | --- | --- |
| Mentors (N=43) | 34 (79.1) | 37 (86.0) | 29 (67.4) |
| Mentees (N=37) | 26 (70.3) | 23 (62.2) | 18 (48.6) |

**Table 3:**
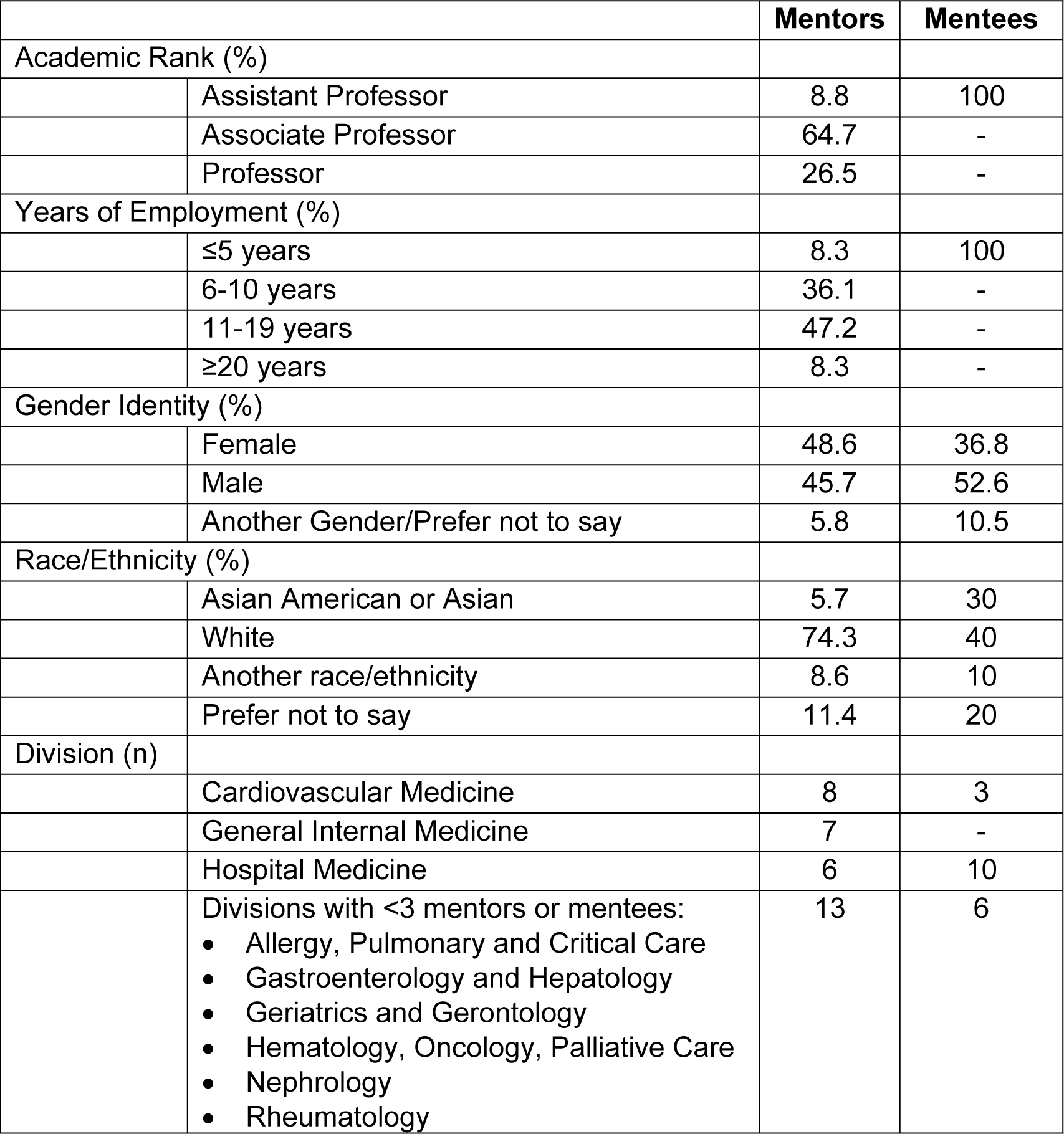
Demographics of Clinical Faculty Mentoring Program Survey Respondents (Post-Survey).

|  |  | **Mentors** | **Mentees** |
| --- | --- | --- | --- |
| Academic Rank (%) |  |  |  |
|  | Assistant Professor | 8.8 | 100 |
|  | Associate Professor | 64.7 | - |
|  | Professor | 26.5 | - |
| Years of Employment (%) |  |  |  |
|  | ≤5 years | 8.3 | 100 |
|  | 6-10 years | 36.1 | - |
|  | 11-19 years | 47.2 | - |
|  | ≥20 years | 8.3 | - |
| Gender Identity (%) |  |  |  |
|  | Female | 48.6 | 36.8 |
|  | Male | 45.7 | 52.6 |
|  | Another Gender/Prefer not to say | 5.8 | 10.5 |
| Race/Ethnicity (%) |  |  |  |
|  | Asian American or Asian | 5.7 | 30 |
|  | White | 74.3 | 40 |
|  | Another race/ethnicity | 8.6 | 10 |
|  | Prefer not to say | 11.4 | 20 |
| Division (n) |  |  |  |
|  | Cardiovascular Medicine | 8 | 3 |
|  | General Internal Medicine | 7 | - |
|  | Hospital Medicine | 6 | 10 |
|  | Divisions with <3 mentors or mentees:<br>• Allergy, Pulmonary and Critical Care<br>• Gastroenterology and Hepatology<br>• Geriatrics and Gerontology<br>• Hematology, Oncology, Palliative Care<br>• Nephrology<br>• Rheumatology | 13 | 6 |

### CFMP Mentor Outcomes

Our primary findings describe changes in mentor job satisfaction, teaching attitudes, mentoring knowledge, and confidence in mentoring and coaching skills from before to after participating in the CFMP (Table 4). We identified statistically significant improvements in almost all domains, with the largest and most consistent improvements in knowledge of mentoring competencies and resources, particularly promotion guidelines and processes, promoting career development through education, promoting career development through opportunities for networking, aligning personal career goals with the UW DOM’s overall goals, and managing Imposter Phenomenon. Absolute improvements in these areas ranged from 0.69-0.88 points (all p<0.001). We also observed statistically significant improvements in confidence in coaching and mentoring skills in each of these areas, though the absolute magnitude of improvement was slightly lower (0.49-0.55 points, all p≤0.02).

**Table 4.** Changes in Mentor Job Satisfaction, Teaching Attitudes, Mentoring Knowledge, and Confidence after Completing the Clinical Faculty Mentoring Program (scale: 1.0 [“not at all] - 5.0 [‘a great deal”]).

|  | Pre-Mean | Standard Deviation | Post-Mean | Standard Deviation | P-Value |
| --- | --- | --- | --- | --- | --- |
| **Job Satisfaction & Attitudes** |  |  |  |  |  |
| I understand my current job in the DOM | 4.55 | 0.69 | 4.72 | 0.53 | 0.202 |
| I am satisfied with the amount of time I have to meet and fulfill the obligations of my job description | 3.25 | 1.11 | 3.79 | 1.05 | **0.017** |
| Overall, I am satisfied with my career | 3.93 | 0.75 | 4.31 | 0.66 | **0.001** |
| I am confident about my path to promotion | 4.00 | 0.74 | 4.64 | 0.49 | **<0.001** |
| My day-to-day activities give me a sense of accomplishment | 3.90 | 0.90 | 4.38 | 0.73 | **0.041** |
| The DOM supports work-life balance | 3.28 | 1.10 | 3.72 | 0.96 | **0.017** |
| I am satisfied with my own work-life balance | 3.17 | 1.10 | 3.55 | 1.12 | **0.046** |
| **Teaching Attitudes** |  |  |  |  |  |
| I am satisfied with the influence I have over the focus of my teaching | 4.28 | 0.75 | 4.59 | 0.64 | 0.148 |
| I am satisfied with the extent that my teaching contributes to promotion | 4.04 | 0.85 | 4.54 | 0.71 | **0.010** |
| I am satisfied with the balance of clinical and teaching duties | 3.76 | 1.09 | 4.25 | 0.80 | **0.020** |
| **Knowledge of Mentoring Competencies and Resources** |  |  |  |  |  |
| Promotion guidelines and process | 2.62 | 0.82 | 3.50 | 1.04 | **<0.001** |
| Promoting career development through education | 2.38 | 0.73 | 3.25 | 0.97 | **<0.001** |
| Promoting career development through opportunities for networking | 2.38 | 0.73 | 3.11 | 0.83 | **<0.001** |
| Aligning personal career goals with DOM's overall goals | 2.55 | 0.87 | 3.25 | 0.93 | **<0.001** |
| Burnout recognition & mitigation | 2.79 | 0.82 | 3.32 | 0.94 | **0.005** |
| Building resilience | 2.66 | 0.72 | 3.25 | 0.93 | **0.001** |
| Supporting diversity, equity, and inclusion in the workplace | 2.83 | 0.89 | 3.39 | 1.03 | **0.007** |
| Managing Imposter Phenomenon | 2.52 | 1.06 | 3.21 | 0.83 | **<0.001** |
| **Confidence in Coaching/Mentoring Skills** |  |  |  |  |  |
| Promotion guidelines and process | 2.72 | 0.80 | 3.21 | 0.83 | **0.002** |
| Promoting career development through education | 2.59 | 0.73 | 3.14 | 0.76 | **<0.001** |
| Promoting career development through opportunities for networking | 2.55 | 0.83 | 3.07 | 0.90 | **0.001** |
| Aligning personal career goals with DOM's overall goals | 2.76 | 0.95 | 3.25 | 0.84 | **0.020** |
| Burnout recognition & mitigation | 2.83 | 1.04 | 3.29 | 0.85 | **0.035** |
| Building resilience | 2.82 | 0.94 | 3.25 | 0.93 | **0.043** |
| Supporting diversity, equity, and inclusion in the workplace | 2.82 | 0.98 | 3.21 | 0.99 | **0.011** |
| Managing Imposter Phenomenon | 2.64 | 1.06 | 3.14 | 1.04 | **<0.001** |

Mentors’ confidence in modeling most coaching behaviors did not change appreciably after participation in mentor training and meeting with their mentees (Figure 2). The two question responses with the strongest evidence for improvement were “Supporting the mentee to integrate new awareness, insight, learning into their worldview and behaviors” (p=0.0503) and “Managing time and focus of mentoring session (p=0.022).” For 9 of 10 questions, the modal response for the post-survey was the same as in the pre-test. Results were similar by gender identity and by years of experience (≤10 vs >10 years). Means of mentors’ perceptions of the UW DOM climate, their professional fit, equitable procedures, and support for professional development were high at baseline (3.90-4.31, on a scale of 1.0 [“not at all] - 5.0 [‘a great deal”]); post-survey mean values increased for these domains, but differences were not statistically significant (data not shown).

**Figure 2.**
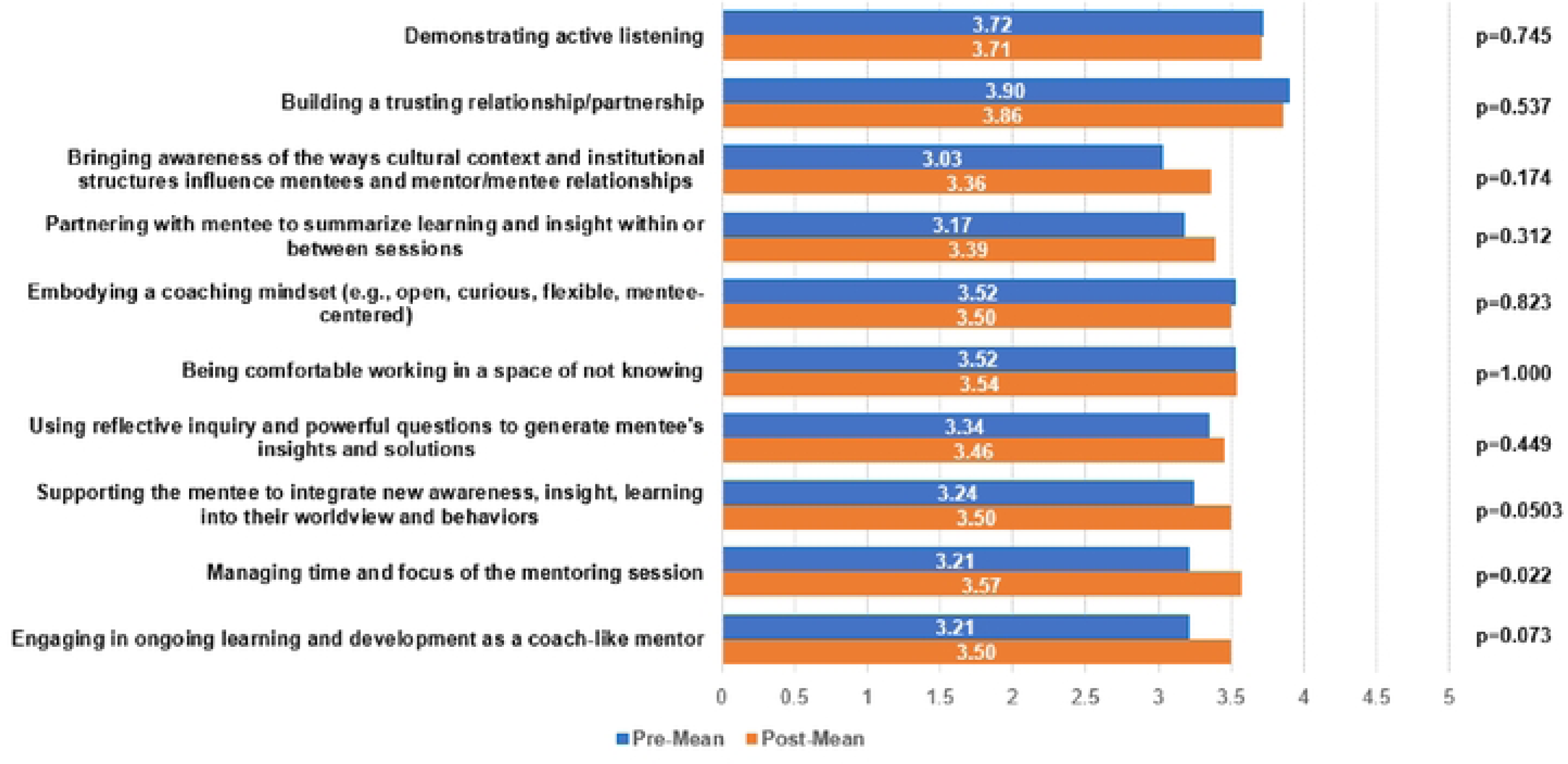
Mentor Changes in Confidence at Modeling Coaching Behaviors (Means, Pre-Post) (scale: 1.0 [“not at all] - 5.0 [’a great deal”]).

Overall, 44% of respondents found the strategies used in the coach approach to mentoring program to be “very helpful” and another 44% found them to be “somewhat helpful” with one respondent selecting “somewhat unhelpful.” Similarly, 56% responded that they were “very likely” to use a coach approach to mentoring junior faculty and 28% responded that they were “somewhat likely” to use this approach, with 8% responding that were “somewhat unlikely” or “unlikely” to use it.

Representative narrative comments from mentors about the aspects of the CFMP are summarized in Table 5. Mentors’ comments highlighted the practical aspects of the program, sense of community, appreciation for the “formal structure and progressive nature of the training,” and the overall effect participation in CFMP had on individual mentoring practices. Some mentors expressed differing views on the coach approach to mentorship. Comments also focused on the challenge of the time commitment necessary for mentor training and mentorship meetings.

**Table 5.**
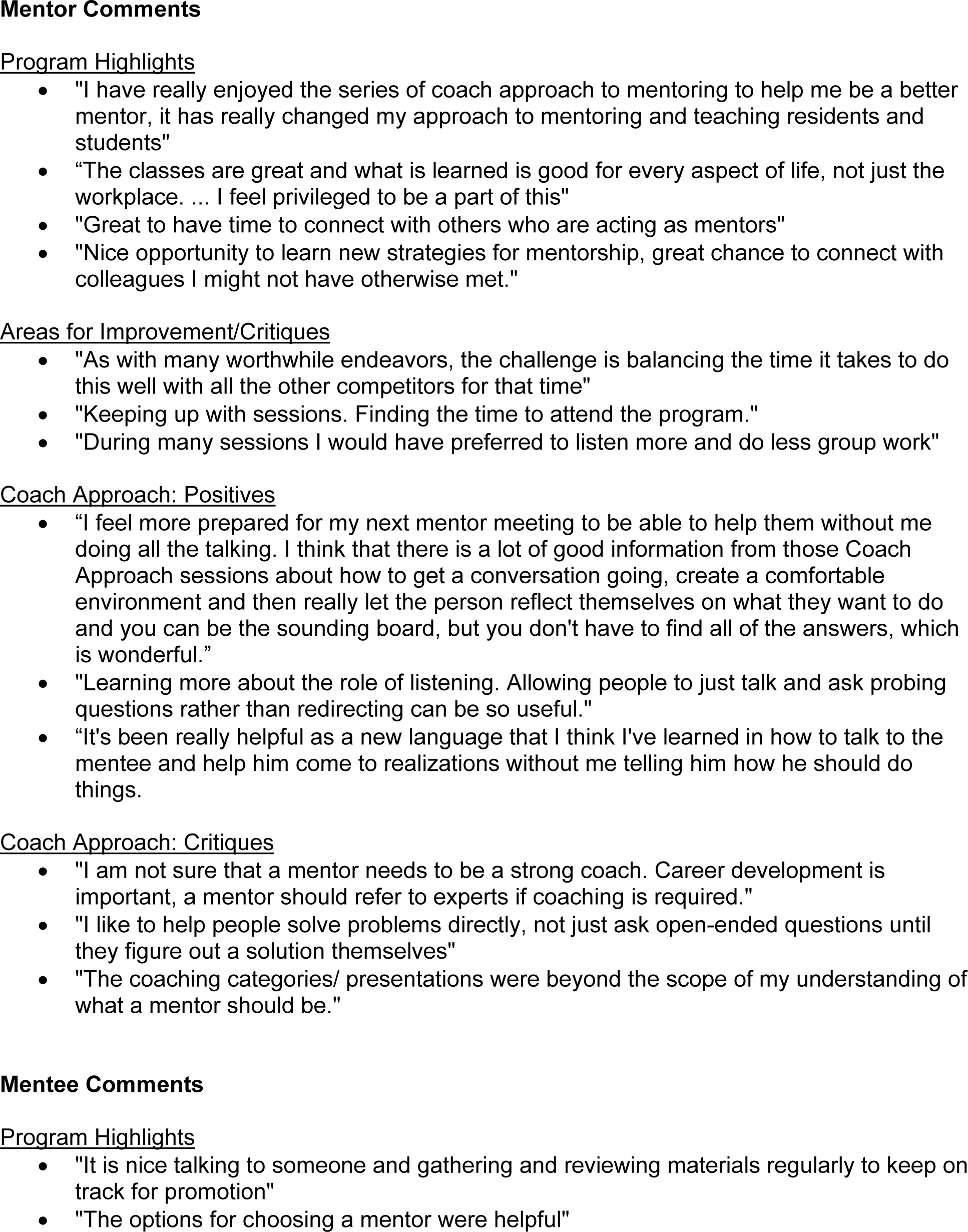
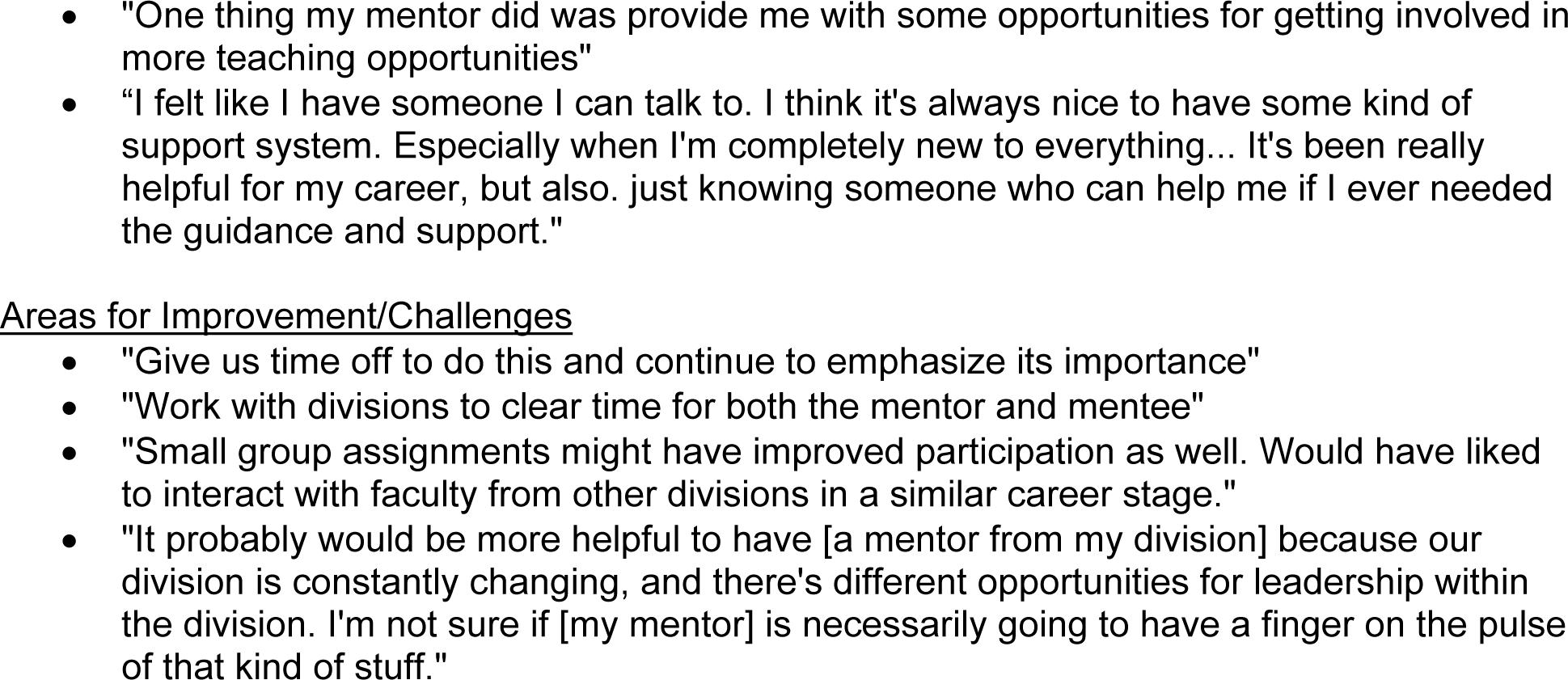
Representative Narrative Comments from Mentor and Mentee Participants in the Clinical Faculty Mentoring Program Mentor Comments.

### CFMP Mentee Outcomes

Surveyed mentees reported high baseline pre- and post-mean levels of agreement on items assessing understanding their current job description, job/career satisfaction, path to promotion, and work-life balance (data not shown). Mentees reported increased confidence in all eight domains of emphasis from mentoring sessions, but pre-post differences in means did not reach statistical significance given the small sample size (Figure 3). Numerically, at least half of participants reported higher confidence on the post-survey than on the pre-survey for all but one of these measures (aligning personal career goals with DOM’s overall goals; data not shown). Representative narrative comments from mentees regarding program highlights and areas for improvement are summarized in Table 5. Qualitative responses echoed a similar theme as that of mentors related to difficulty with finding time for preparing for and participating in mentoring meetings.

**Figure 3.**
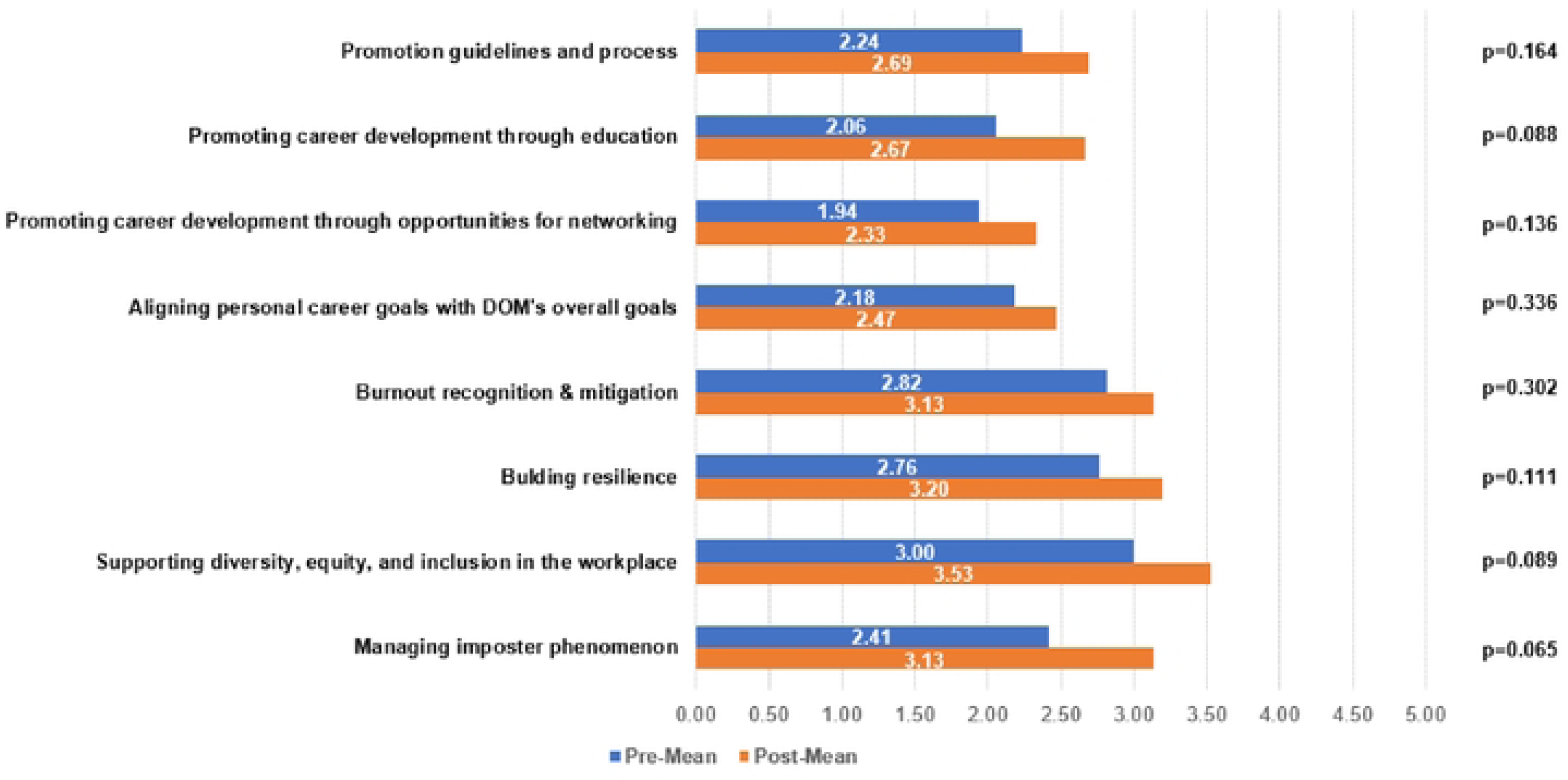
Mentee Changes in Skills and Knowledge (Means, Pre-Post) (scale: 1.0 [“not at all] - 5.0[’a great deal”]).

### CFDS Outcomes

From September 2022 through June 2023, 39 sessions that were attended by 239 unique faculty. On average, 38 individuals participated in each session (range 22-59); 44% were Assistant Professors, 34% were Associate Professors, and 22% were Professors. The sessions with the highest attendance were on the topics of time management and organization skills (n=50), promotion (49), physician burnout (47), being a woman in medicine (47), building an anti-racist environment (45), feeling fulfilled by your job (43), learning climate (43), promoting clinical reasoning (43), setting professional goals (42), and public speaking (42).

The post-CFDS survey was completed by 99 faculty. Respondents represented all 11 Divisions in the UW DOM. Overall responses were favorable with 70% of respondents agreeing or strongly agreeing that the skills they learned helped them professionally and 56-60% agreeing or strongly agreeing that they learned to be a better teacher in the clinical environment, learned how to recognize and address bias, and that the skills they learned are important for leadership. Positive themes that emerged from narrative comments were appreciation for the variety of topics, interactive format, and opportunity to gain perspectives from other faculty. The sessions on clinical teaching were particularly well-received. The most notable challenge was the timing of the sessions given the busy schedules of clinical faculty.

## Discussion

The UW DOM Clinical Faculty Development Program combined a novel professional coach approach to formal mentoring (the CFMP) and a recurring series of interactive sessions aimed at building a diverse set of skills and boosting the vitality of its clinical faculty (the CFDS). Our primary finding was that mentors who participated in the CFMP reported statistically significant improvements in almost all domains queried with notable self-reported improvements in knowledge of and confidence in using coaching skills to mentor junior faculty in the areas of promotion guidelines and processes, promoting career development through education, promoting career development through opportunities for networking, aligning personal career goals with the DOM’s overall goals, and managing Imposter Phenomenon. The vast majority of mentor respondents (88%) found the strategies used in the coach approach to mentoring program to be “very” or “somewhat” helpful, 84% responded that they were “very” or “somewhat likely” to use a coach approach to mentoring junior faculty, and 28% responded that they were “somewhat” likely to use this approach when mentoring junior faculty. All mentees had at least one meeting with a mentor and mentees reported increased confidence in each domain we focused on. The CFDS was well-attended and appeared to support the outcomes from the CFMP by fostering a culture of mentorship, facilitating professional skills-building, and providing opportunities for interpersonal interactions.

Mentoring programs at academic health centers often are project-focused and outcome-driven^5,7,12^. Although we used a dyad mentorship model and adhered to best practices in the science of effective mentoring, our CFMP was novel by using coaching as the basis of mentor training, rather than the traditional “mentor as problem-solver and advisor” approach. Coaching aims to enhance mentee’s self-awareness and growth and is based on principles from positive psychology and motivational interviewing^20^. Professional development coaching programs in health professions have beneficial effects such as aiding faculty in achieving professional goals, decreasing reports of burnout, and increasing work engagement and satisfaction^21,22^. Coaching is a skill that clinical faculty can apply in leadership roles and when working with learners and advanced practice providers^23,24^. After coach training, mentors in the CFMP felt the coaching skills were effective and impactful for mentoring junior faculty. To understand the extent to which mentors internalized and implemented behaviors taught in the coach approach training, we assessed their level of confidence to model specific coaching behavior. Most domains showed only small improvements when assessed in aggregate (*i.e.,* group means) or using individual change scores. However, the two domains showing greater evidence of change - (i) supporting the mentee to integrate new awareness, insight, learning into their worldview and behaviors and (ii) managing time and focus of mentoring sessions - are two of the more critical aspects of what makes a coach approach such a powerful paradigm for mentors. Coaching, at its root, is about creating deeper learning that promotes action to create behavioral change. Being able to coach a mentee to successfully integrate new awareness into their worldview and to act accordingly, coupled with the very practical skill of managing time and focus of a mentor coaching session, are keys to the overall success of any coaching or coach-approach engagement. Improving confidence in these two coaching behaviors positions the mentor/mentee relationship for success. We also suspect that these trends reflected “response shift bias,” whereby mentors “did not know what they did not know” in the pre-survey; that is, as they took part in mentoring throughout the year, they recalibrated their confidence based on what they learned. Indeed, several participants indicated this informally during the training sessions. Other faculty mentoring programs also have described differences between mentor confidence in mentoring competencies and adopting mentoring behaviors ^25^. These findings highlight the need not only for effective mentor training but also for follow-up and continuous mentorship skills building.

The CFMP was initiated at the same time as the CFDS. The CFDS was open to all clinical faculty in the UW DOM, so these activities overlapped temporally and many CFMP participants attended CFDS sessions Thus, observations about the impact of CFMP must be considered on the background of this program. In addition to its unique focus on faculty development topics of specific interest to clinicians, the CFDS supported the outcomes of the CFMP due to overlapping topics and exposure of CFMP participants to presenters and participants from outside the program. Another unique aspect of both the CFMP and CFDS was that they were initiated and conducted virtually due to the COVID-19 pandemic. Virtual sessions permitted a wider audience to attend, including faculty working at distant clinical sites and those working from home, as observed in another study^26^. Furthermore, digital recordings allowed asynchronous viewing for faculty. The chat function and breakout rooms were options for greater audience participation, creating a more active learning environment. Virtual presentations were well received, though some faculty still expressed a desire for face-to-face experiences.

Finding time to participate in the CFMP and CFDS was the major challenge identified by respondents. Faculty in the CFMP agreed that the time reserved for mentor training and annual meetings was fair and sufficient; however, finding time in a busy clinician’s day for ongoing mentor-mentee meetings and participation in the noon-hour CFDS sessions were challenged by competing clinical and administrative demands. Addressing time constraints would allow faculty to engage more consistently, further cultivating a vibrant culture of faculty development. Future directions to assure sustainability include increasing the pool of mentors and opportunities for mentor trainees, creating digital tools to facilitate implementation of mentoring strategies and other skills, and central administrative support for scheduling meetings.

### Limitations

Since our program enrolled early career faculty who mostly were in their first year of employment and it initiated during a late stage of the pandemic, we do not know the impact of these programs on longer-term outcomes such as promotion, teaching effectiveness, wellbeing, and faculty vitality. Success in promotion and scholarship from faculty participating in faculty development programs typically have been measured 3-10 years after program initiation^5,6,12^. However, there were signals in our data that faculty participation in these programs improved confidence, work satisfaction, and perceptions of the DOM’s support of their professional development and mentorship, even in the first year with most mentees having only 1-2 sessions with their mentors.

In addition to response shift bias, a major limitation of our data is response bias. The data presented reflect the responses of people who participated and responded to the pre- and post-surveys. We do not know the responses of those who chose not to respond. Because the survey responses were confidential, we were not able to identify and contact participants who did not respond, and thus could not provide more detailed data analyses by race/ethnicity or academic division due to the small numbers of participants in those subgroups.

## Conclusions

The UW DOM Clinical Faculty Development Program created a novel, coach approach to mentoring and a weekly faculty development series to provide clinicians with experiences that promoted skill-building, mentorship, and professional opportunities. Mentors who participated in the CFMP reported improved knowledge of and confidence in using coaching skills to mentor junior faculty in promotion guidelines and processes, career development through education and opportunities for networking, aligning personal career goals with our department’s goals, and managing Imposter Phenomenon. Mentors overwhelmingly felt that the program was helpful and will be useful. The CFDS was well-attended and supported the outcomes from the CFMP. Longitudinal follow-up is needed to determine how this program affects mentees and if it achieves its long-term goals.

## Declarations

### Ethics approval and consent to participate

The UW Health Sciences Human Subjects Committee determined that our evaluation of UW CFMP and CFDS did not meet the definition of human subjects research. They determined that the activities and analyses described in this report were considered quality assurance and declined to review them. Participation was voluntary and part of a quality review project. Since our IRB declined to review it, we were not able to obtain valid informed consent.

### Consent for publication

Not applicable.

### Availability for data and materials

The datasets generated and/or analysed during the current study are not publicly available due personally identifying information but are available from the corresponding author on reasonable request.

### Author contributions

- James D Alstott - interpretation of data, drafted manuscript
- Chariti Gent - conception and design, interpretation of data, critical revision
- Christine Fabian Bell – analysis and interpretation of data, critical revision
- Daniel R Marlin – analysis and interpretation of data, critical revision
- Anthony Hernandez – design, critical revision
- Esther Schulman - conception and design, interpretation of data, critical revision
- Sharon Gehl - conception and design, interpretation of data, critical revision
- Lynn M Schnapp - conception and design, interpretation of data, critical revision
- James H Stein – conception and design, interpretation of data, critical revision All authors read and approved the final manuscript.

## Data Availability

Our data can be shared by request tot he corresponding author, however UW rule prohibit disclosure of information that might identify individual participants.

## Acknowledgements

Not applicable.

## Abbreviations

CFDS: Clinical Faculty Development Series
CFMP: Clinical Faculty Mentoring Program
CT: Clinician-Teacher
DEI: Diversity, Equity and Inclusion
DOM: Department of Medicine
ICF: International Coaching Federation
URiM: UnderRepresented in Medicine
UW: University of Wisconsin

